# Concordance of high blood pressure among middle-aged and older same-sex couples in the USA

**DOI:** 10.1101/2024.01.09.24300695

**Authors:** Jithin Sam Varghese, Michael G. Curtis, Samuel C.O. Opara, Shivani A. Patel, Anandi N. Sheth, Sophia A. Hussen

## Abstract

Heterosexual couples in romantic relationships are known to influence each other’s hypertension risk. However, the role of partners on an individual’s hypertension status in same-sex relationships is less understood. Our objective is to characterize the burden of high blood pressure among middle-aged and older couples consisting of men who have sex with men (MSM) and women who have sex with women (WSW) living in the US. Among 81 same-sex couples from the Health and Retirement Study 2006-18, 72.4% (95%CI: 23.4-95.7) MSM couples and 61.0% (95%CI: 30.4-84.8) WSW couples consisted of both partners with hypertension. Same-sex couples demonstrate high concordance of hypertension and related risk factors, suggesting a need to develop novel interventions targeting MSM and WSW couples

## Introduction

Hypertension affects 115 million adults in the United States (US).^1^ Heterosexual couples in romantic relationships are known to influence each other’s hypertension risk (including overweight/obesity) and hypertension status, suggesting couple-focused interventions may be beneficial for long-term disease management.^2^ However, the role of partners on an individual’s hypertension status in same-sex relationships is less understood, despite substantial health disparities for sexual minorities relative to the general population.^3,4^ Our objective is to characterize the burden of overweight/obesity and high blood pressure among middle-aged and older couples consisting of men who have sex with men (MSM) and women who have sex with women (WSW) living in the US.

## Methods

We used data from the Health and Retirement Study (2006-2018), which contains nationally representative, longitudinal household surveys of individuals aged 50 years and older, and their partners. Couples were defined as same-sex individuals who self-reported being in a romantic relationship with each other. Gender and sexual orientation were not independently ascertained. The analytic sample consisted of 39 MSM and 42 WSW couples, among whom both partners had measured blood pressure.

We conducted a pooled analysis of seven study waves of the HRS, given the limitations in sample size, separately for MSM and WSW couples. We used the latest available data on hypertension for each primary respondent and their partner. Blood pressure was measured thrice for each respondent using validated electronic monitors, and the average of the last two measurements were used to estimate systolic and diastolic blood pressure. Participants were considered as having high blood pressure if they met one of the following criteria: history of self-reported high blood pressure (or hypertension) based on a provider diagnosis, systolic blood pressure ≥ 130 mmHg or diastolic blood pressure ≥ 80 mmHg.^1^ Overweight or obesity was defined as body mass index ≥ 25.0 kg/m^2^, derived from height and weight measured by trained enumerators using standardized instruments. All analysis considered the complex survey design and applied sample weights provided. We compared these estimates with those from heterosexual couples surveyed as part of the Health and Retirement Study in 2016.^2^

## Results

Among same-sex couples drawn from nationally representative surveys of adults 50 years or older in the US, 79.4% (31 out of 39) and 71.4% (30 out of 42) were measured in 2016 or later for couples comprising men who have sex with men (MSM) and women who have sex with women (WSW), respectively. Only 1.3% (95%CI: 0.4-3.9) of MSM but 53.1% (95%CI: 23.3-80.9) of WSW were married. At least one partner was 65 years or older among 64.5% (95%CI: 21.3-92.4) MSM couples and 32.7% (95%CI: 11.8-63.7) WSW couples. The majority of couples consisted of non-Hispanic White partners among both MSM (80.8% [95%CI: 42.8-96.0]) and WSW (92.6% [95%CI: 68.5-98.6]).

Both partners had overweight or obesity in 64.2% (95%CI: 21.2-92.3) MSM couples and 35.6% (95%CI: 13.8-65.6) WSW couples. Similarly, 72.4% (95%CI: 23.4-95.7) MSM couples and 61.0% (95%CI: 30.4-84.8) WSW couples consisted of both partners with hypertension (**Figure**). The burden of concordant hypertension is likely similar or higher than heterosexual couples (54.3% [95%CI: 52.2-56.3]) surveyed in 2016, although estimates for same-sex couples have low precision (**Figure**).

**Figure.**
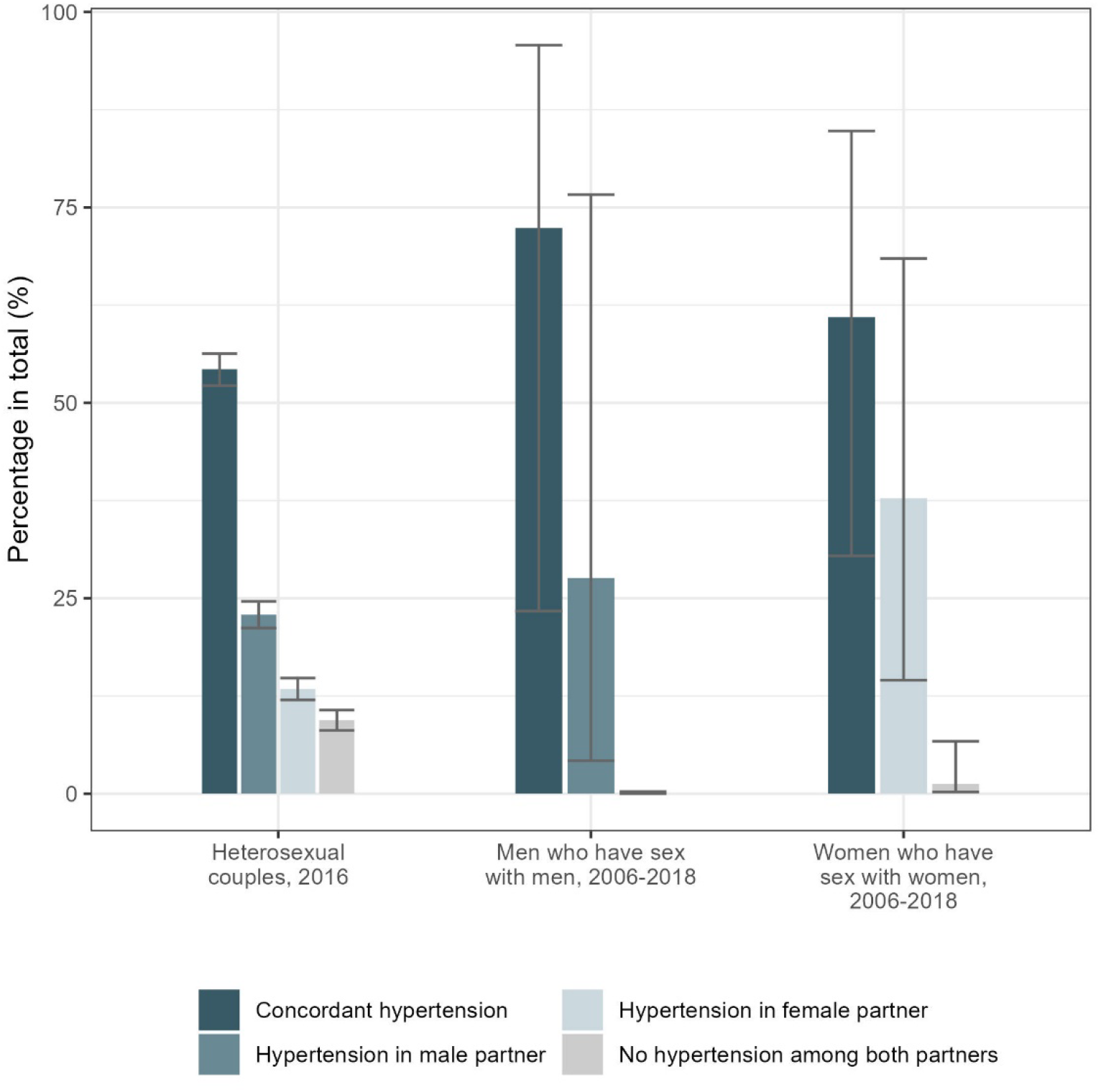
Concordance of high blood pressure among heterosexual and same-sex couples in Health and Retirement Study. All estimates are survey weighted proportions and 95% confidence intervals. We first normalized sample weights for an observation by dividing it by the sum of sample weights for the wave. Next, we inverse-weighted the normalized sample weight with proportion of participants in the pooled serial cross-sectional dataset corresponding to that wave to account for wave-specific sample sizes. Mean (SD) age for heterosexual wives and husbands were 62.9 (9.7) years and 65.7 (9.7) years respectively. Mean (SD) age for the younger and older partner in MSM couples (n = 39) were 53.9 (10.1) years and 61.4 (5.4) years. Mean (SD) age for the younger and older partners in WSW couples (n = 42) were 54.5 (5.2) years and 61.4 (5.9) years.

## Discussion

Using nationally representative data from 2006 to 2018, this exploratory study demonstrated a high burden of concordant hypertension among middle-aged and older same-sex couples living in the US. Concordance rates may be similar or higher in same-sex than in heterosexual couples.^2^ The majority of MSM and WSW couples also demonstrated concordance in overweight or obesity. These results suggest that couple-centered strategies designed for hypertension and related cardiovascular risk management in heterosexual couples may be particularly beneficial for same-sex couples.

Limitations of this study include pooling data over 12 years due to paucity of data. We did not explore if concordance was beyond what is statistically expected by chance and were unable to examine racial-ethnic differences due to underrepresentation of racial/ethnic minority groups. Additionally, although the estimates of same-sex marriage rates in this study (56.3%) were similar to what is observed nationally for all WSW couples (58.6%), the rates of marriage for MSM couples are much lower in this study (1.6% versus 57.3% nationally).^5^ This requires further exploration and suggests possible non-representativeness of MSM couples included. Finally, although the 2017 ACC/AHA guidelines recommend diagnosis of hypertension based on ≥ 2 readings taken on ≥ 2 visits, only blood pressure measurements from the latest survey were included.^1^

In summary, same-sex couples demonstrate high concordance of hypertension and related risk factors, suggesting a need to develop novel interventions targeting MSM and WSW couples. The small sample size of this exploratory study also highlights the need to study couple concordance further in sexual minority couples. Future studies of partner influences on health among same-sex couples should be conducted, especially among racial-ethnic minorities who have disproportionately high burden of hypertension.

## Abbreviations

ACC/AHA: American College of Cardiology/American Heart Association
ISR: Institute for Social Research
MSM: Men who have sex with men
WSW: Women who have sex with women

## ACKNOWLEDGEMENT/SUPPORT

### Ethics approval and consent to participate

All participants gave written informed consent before participation. We were exempted from ethical approval for this secondary data analysis from the Institutional Review Board of Emory University.

### Data Availability Statement

All datasets used in this analysis are available for download at https://hrs.isr.umich.edu. The code for the analysis is available on https://github.com/jvargh7/msm_hiv_htn/tree/main/hrs.

### Consent for publication

Not applicable

### Funding

Emory Global Diabetes Research Center. Michael G. Curtis was supported by National Institute of Allergy and Infectious Diseases (T32AI157855) and Emory Center for AIDS Research (P30AI050409). Shivani A. Patel was supported by National Heart, Lung and Blood Institute (P01HL154996-01A1).

### Author contributions

JSV and SH developed the study and the analysis plan with inputs from all authors. JSV had full access to the data in the study and takes responsibility for the integrity of the data and the accuracy of the data analysis. JSV wrote the first draft with inputs from SH. All authors edited and approved the manuscript.

## Acknowledgements

We thank the participants and survey enumerators of Health and Retirement Study 2006-2018.

